# How does corruption influence health system efficiency? A case study of two counties in Kenya

**DOI:** 10.1101/2023.06.27.23291949

**Authors:** Joshua Munywoki, Lizah Nyawira, Anita Musiega, Rebecca G Njuguna, Benjamin Tsofa, Kara Hanson, Sassy Molyneux, Charles Normand, Julie Jemutai, Edwine Barasa

## Abstract

**Objective:** Efficiency gains are one potential pathway to unlocking additional resources for the health sector. Given that corruption has been cited as a key cause of inefficiency in the health sector, the objective of this study was to examine the influence of corruption on the efficiency of county health systems in Kenya.

**Design, setting and analysis:** We conducted a qualitative case study in two counties in Kenya. We developed a conceptual framework from a literature review to guide the development of tools and analysis. We collected qualitative data through in-depth interviews (n=26) with county, sub-county, and health facility level respondents across the two counties. We analyzed the data collected using a framework approach.

**Results:** Corrupt practices reported in the case study counties included non-merit-based recruitment and training of health workers, supply of substandard goods, equipment and infrastructure; theft, embezzlement and misuse of public funds and property; and informal payments. These practices were perceived to impact negatively on health system efficiency by leading to a direct loss of health sector resources, increase in operational costs, poor quality of care, reduced staff motivation and productivity, and reduced access to healthcare services.

**Conclusion:** The efficiency of county health systems could be enhanced by implementing anti-corruption strategies to tackle the identified corrupt practices.

**Strengths and limitations of this study:**

- To the best of our knowledge, this paper is the first of its kind to focus on how corruption affects attainment of health sector goals in Kenya
- Our reported findings focus on only two out of forty-seven counties in Kenya. Transferability of our findings ought to be interpreted with consideration to contextual factors that shaped reported corruption practices
- We found study respondents to be cautious when reporting their experiences and perceptions on health sector corruption. It is possible that social desirability bias affected response from our study participants
- Given the sensitivity and difficulty of corruption as a study topic, this study was not able to evaluate the effectiveness of existing anti-corruption strategies in Kenya

## INTRODUCTION

Kenya has made a commitment to achieve UHC by 2030 (1). The goal of UHC is for everyone to have access to quality preventive, promotive, curative, and palliative health care, without getting into financial difficulties (2). Kenya’s progress towards UHC is constrained by resource scarcity. While it is recommended that LMIC public spending on healthcare be at least 5% of gross domestic product (GDP) (3), Kenya ’s public health spending as a share of GDP is 2% (4). In addition to allocating more resources to the health sector, additional resources for the health sector could potentially be unlocked by efficiency gains (5). Health system efficiency refers to the maximization of health goals given the available resources (6). It has been estimated that 20-40% of healthcare resources are lost due to inefficiency (7).

Corruption has been cited as one of the major sources of inefficiency in the health sector (8). Corruption is defined as the misuse of entrusted power for private gain (9). Corruption occurs when public officials who have been given the authority to carry out goals which further the public good, instead use their position and power to benefit themselves and others close to them (9). It is estimated that between 10% and 25% of global spending on public procurement of health is lost through corruption (10). Corruption thus results in loss of health system resources and undermines operational efficiency of the health system by increasing the cost of health services, reducing the quality of services provided and increasing inequity in access to health services by disadvantaging the economically vulnerable (11–15). Corruption as an issue affecting the health system has, however, been scarcely studied (9,16,17). This is partly because corruption is perceived by both policy makers and researchers to be a difficult and sensitive topic to discuss (18,19). There is thus scarcity of research evidence on the effects of corruption on health systems (9,16,18,20,21).

Kenya has a devolved system of government, with a national government and 47 semi-autonomous county governments (22). The health sector is the most devolved sector, with the national government retaining policy and regulatory functions, while the county governments are responsible for service delivery (23). County governments are funded by a grant allocated by the national government, donor funds, and locally generated revenues (24). Counties develop budgets and allocate funds across sectors. In the health sector, counties recruit and manage health workers, manage procurement, and own and manage public healthcare facilities (23,25).

This study focuses on corruption at the county health system in the Kenya health sector. It is part of a larger study, the Kenyan Efficiency Study (KES), that aimed to examine the level and determinants of health system efficiency at the county level in Kenya. The first phase of the study, that engaged stakeholders in the Kenyan health sector to explore their perceptions about factors that affect the efficiency of county health system, highlighted corruption as one of the factors (26). The second phase of the study measured relative efficiency of the 47 county health systems and identified relatively efficient and inefficient counties (27). This paper reports findings of the third phase of the study that aimed to conduct county case studies on selected factors. Specifically, this paper examines views on the influence of corruption on the efficiency of county health systems in Kenya.

## METHODS

### Conceptual Framework

Figure 1 outlines the conceptual framework that we developed drawing from literature we reviewed on the common forms of corruption in the health sector (9,28). The conceptual framework focuses on health sector corruption associated with human resource management, procurement, theft, embezzlement, and misuse of public resources and property, and informal healthcare payments. The framework proposes that these forms of corruption influence the efficiency of health systems by causing resource losses, increasing operational costs of the health sector, reducing the motivation and productivity of the health workforce, reducing quality of care, and reducing access to healthcare services.

**Figure 1:**
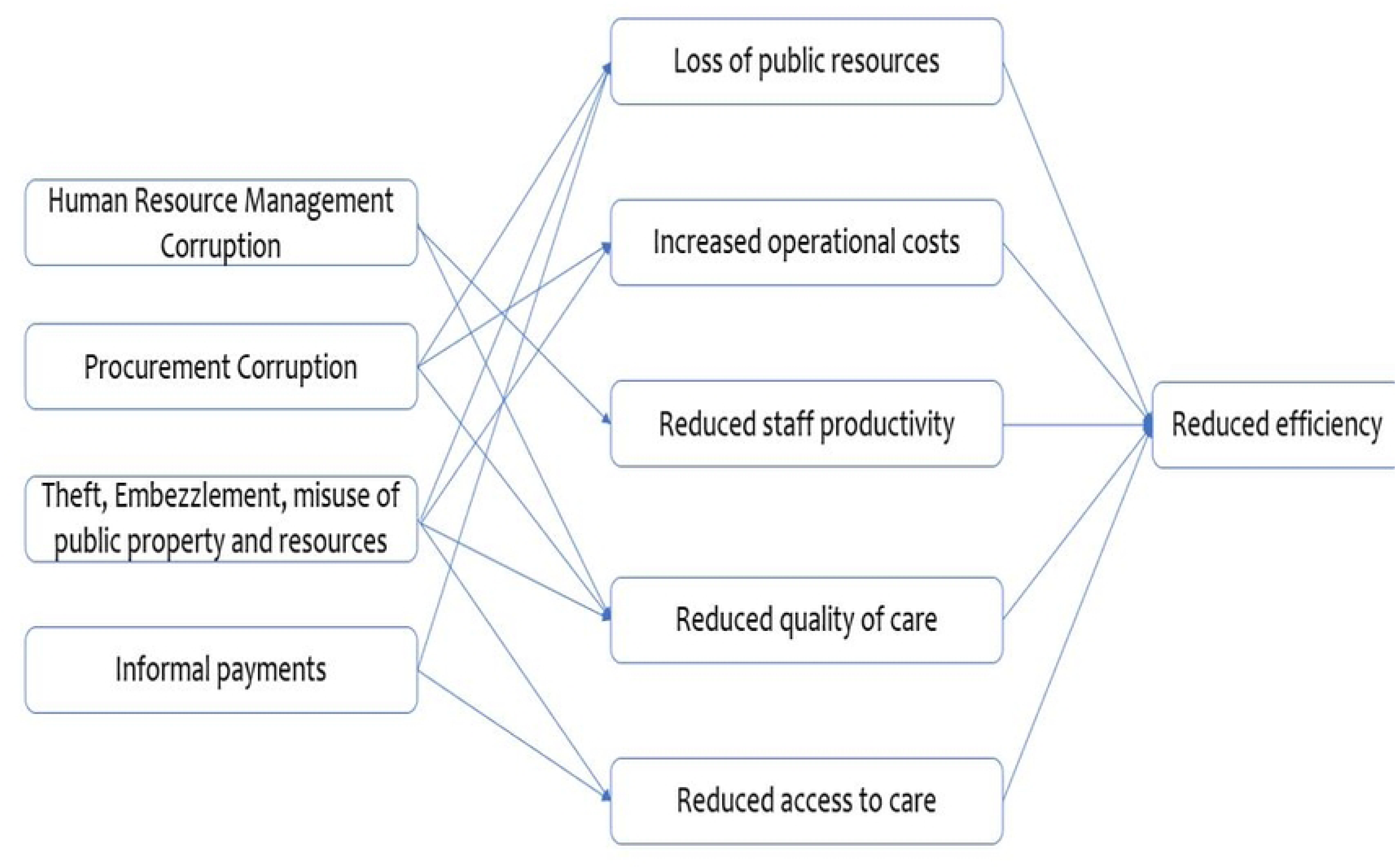
Study conceptual framework.

### Study Design

We used a qualitative case study approach using county health systems as our cases. We selected two counties purposefully based on relative efficiency scores that were computed in the second phase of the larger, Kenya Efficiency Study (27). We anonymized the counties to minimize the risk of identifying study respondents.

### Study Setting

We collected data at the county level in Kenya from two counties. County A had a high population and density. This county had an efficiency score of 81% (27). County B had a low population and population density. This county had an efficiency score of 52% (27).

### Population and Data Collection

We collected data using in-depth interviews in May and June 2021. In each county, we collected data at the county administration level and at the health facility level. At the county administration level, we carried out interviews with study respondents drawn from the county and sub-county health management teams. At the health facility level, we carried out interviews with respondents drawn from public hospitals, and public primary healthcare (PHC) facilities – Health centres and dispensaries. We selected two public healthcare facilities, a hospital, and a primary health facility in each of the study counties. We selected the individual facilities within the counties through random sampling from the Kenya Master Health Facility List (KHFML).

### In-depth interviews

We carried out purposive sampling of participants for in-depth interviews. We also used a snowballing technique to identify additional participants. We stopped data collection where we felt we had reached data saturation (that is, the point where no new information was obtained with new interviews). We conducted a total of 26 interviews. Table 1 outlines the distribution of study respondents across the levels of the health system and study counties.

**Table 1:**
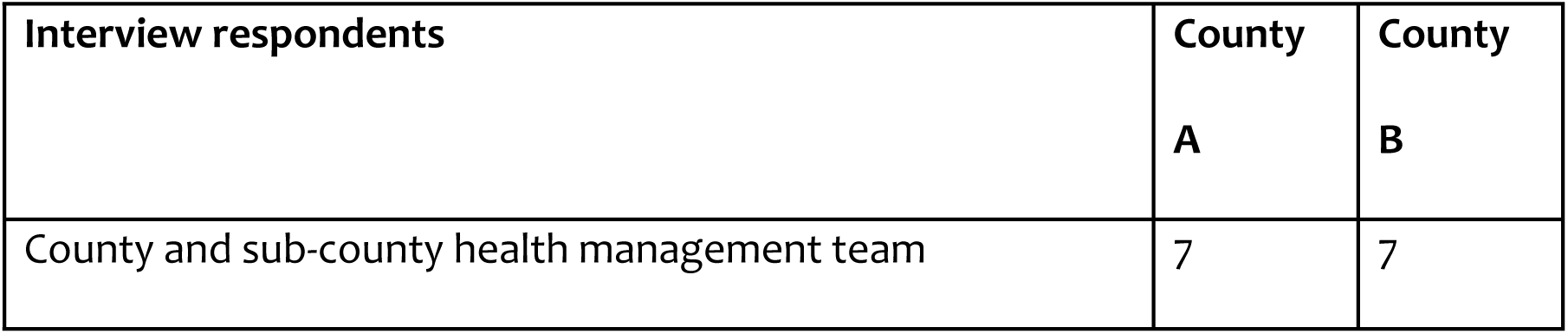
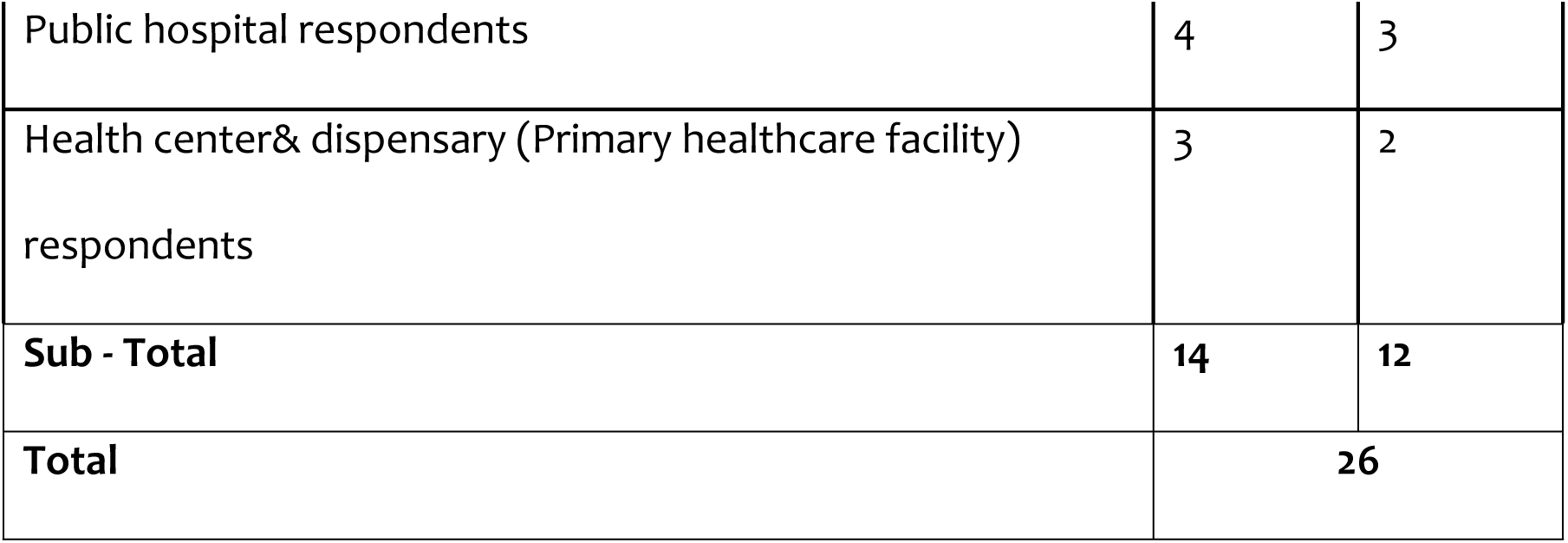
Summary of interview respondents.

We conducted the interviews using a topic guide that was informed by the study’s conceptual framework. Interviews were held at private locations in the working stations of the study respondents or an alternative location that the respondents deemed suitable and confidential. Each in-depth interview lasted approximately 45 to 60 minutes. We audio-recorded all interviews using a digital recorder. We carried out peer debriefing among the researchers to enhance the credibility of the data collected.

### Patient and public involvement

None

### Data management and analysis

We transcribed the qualitative interview audio files into MS Word. We then then cross-checked the transcripts against the audio recordings as a quality assurance measure. We imported the transcripts into NVIVO 10 software (QSR International, Australia) for analysis using a modified framework approach (29). This entailed the development of a coding framework guided by the study’s conceptual framework and the themes emerging from the data. We then coded the transcripts using this coding framework while allowing for the emergence of new themes. We subsequently charted the coded data and synthesized the charted data to develop result narratives that were supported by illustrative quotes.

### Ethical considerations

We obtained ethics approval (KEMRI/SERU/CGMR-C/154/3814) from the KEMRI Scientific and Ethics Review Unit (SERU). We also obtained approvals from other relevant authorities prior to commencement of the study. All study participants provided written informed consent to participating in the study and being audio recorded. We ensured confidentiality by anonymizing the study counties, de—identifying respondent data, securing the collected data in password protected computers, and restricting access to the data to researchers only.

## RESULTS

In this section, we present our findings on how corruption affects the efficiency of county health systems. These findings are presented according to the main forms of corruption identified in the study’s conceptual framework, namely: Human resource management, procurement, theft or misuse of public property, fraud, and embezzlement of funds.

### Corruption In Human Resource Management Processes

#### Non-merit-based recruitment, promotion, and award of training opportunities

Study respondents reported that the recruitment and promotion of staff by the county was not always based on merit. Staff recruitment and promotions were sometimes influenced by bribes and political patronage. The latter was partly possible because of informal arrangements in which local political leaders (Members of the County Assembly) were involved in the county staff recruitment process, opening the process to political influence.

> *“Politicians are allocated a number of staff to be employed…This now affects those who are poor or those who lack a second (influential) person to maybe to push for him or her. These ones lose chances. So the chances just gone to those who know them (politicians) rather than fairly being interviewed (and) passing. No interview (outcome) is followed.”* County Manager 1, County B

It was also reported that favouritism influenced the selection of staff to undergo in-service training. This was thought to happen because of interests in benefiting from allowance (per diems) when undertaking these trainings.

> *“…occasionally the people who sit at the headquarter are the ones who benefit from all the training. Interestingly, some of this training, which is done by the partners, uh, have some form of remuneration. So, everybody wants to have that money, the allowance that is been paid. Or you will see heads of department always running around spending three, four weeks out of the station attending.”* County Manager 7, County B

Non-merit-based HRH recruitment negatively impacted county health system efficiency by leading to the appointment of individuals with inadequate capacity to undertake their roles, leading to sub-standard outputs.

> *“Some recruited staff in most counties lack capacity, to be honest. The reason is because of political appointment. Politicians appoint them when they know very well that they have no capacity.”* County Manager 7, County B

> *“Political influence is the recruitment of staff affects even the quality of the workers that we get. It affects performance because at times we don’t get the people that you as a qualified officer thought would be best for you. You get some that might not deliver.”* County Manager 1, County A

Non-merit-based recruitment and promotion of staff was also reported to affect the motivation of staff who had either been recruited on merit or passed over for promotions that they felt they deserved.

> *“Then, of course, these political hires and promotions demotivate those staff that are hardworking.”* County Manager 5, County A

> *“…mtu hawezi ji promote [someone cannot promote themselves-]. I know I got into the service, through the right way, and my details, are with my employer. So, in the spirit of their honesty, I don’t expect to grease anyone in the office to get what eh, legally or due fully I’m entitled for. So, nimeona hazitendeki, na naona [I have seen promotions are not happening] it’s because of eh, I might not say it’s the health department, rather the county government if I generalize.”* County Manager 8, County B

Interview respondents also reported that non-merit-based HRH recruitments led to misalignment of skillsets of recruited staff and skill needs of the county health department.

> *“Like employment, of course they employed people for their political affiliation…Like for example you are employed like a public health officer, but you never stepped in a class of public health… When they talk about bloated work force, go, and do analysis and you will find that there are people on payroll who do not deserve being on it. But when they do head count, they are the first ones to be counted.”* County Manager 2, County A

The non-merit-based HRH also contributed to insubordination of HRH managers, owing to political protection from their benefactors. Respondents reported that staff recruited because of political connections could not be disciplined or sanctioned because of their political protection. This made it difficult for managers to manage the productivity of their staff.

> *“When somebody has a powerful ‘godfather’, managing that person becomes difficult. Such a person does not take advice or instructions. At the same time, you do not have power to discipline them because of their political protection.” County Manager 2, County A*
>
> *“A manager would write several warning letters to some staff that are not performing to expectations, but nothing happens. The manager cannot take any action because the staff has political connections.? So, it puts you, you know, as a middle manager, in a difficult position. It makes you feel helpless. So why am I bothering myself? Let, whatever happens, happen.”* County Manager 7, County B

### Corruption in Procurement Processes

#### Supply of substandard goods, equipment and infrastructure

Respondents associated supply of substandard goods, equipment, and infrastructure with corruption. It was reported that private suppliers would collude with senior county government officials to get paid for these substandard projects.

> *“The fence is done, eh, substandard. And then somebody says imeagushwa na, na, ndovu [it has been brought down by elephants] and you know, hiyo kitu, ata upepo kidogo ikikuha itaanguka, hakuna kitu imewekwa [that thing, even with a small wind it’s going to come down, because there is nothing there]. And after completion of substandard projects, they get clearance for payment of services provided.”* County Manager 7, County B

Supply of substandard commodities and equipment affected health system efficiency in two ways. Firstly, the use of bribes to win tenders limited resources available to implement intended projects and contributed to substandard outputs.

> *“It (corruption) is always a challenge [laughs]. You know, the, the, the moment you, the moment people put kickback (bribe) first, then the…the merit, you know it delays everything then you don’t get standard. We don’t get the best of this. So of course, corruption is, is a instantly, it’s going to be a big challenge.”* County Manager 5, County B

> *“You are supposed to build a maternity and the rooms are supposed to be of a standard size. But if you are going to reduce the funds, the contractor is going to reduce the size of that building. Because the amount of money, which is given is less because there’s a kickback. So that is not going to be maternity. It is going to be, something like you know, it’s a, it’s a something that looks like maternity. [laughter].”* County Manager 7, County B

Secondly, corruption in tender award processes increased the cost of development projects. For instance, an inefficiently managed implementation project would be allocated additional funds in subsequent years, increasing the costs of its development while limiting funds available for other development projects.

Interviewees reported that corrupt suppliers got away with their malpractices by bribing persons conducting audit queries or using political connectedness to influence auditor reports.

> *“I think our audit system is toothless to be honest with you at the county level, there’s nothing. And I’ve said that in the meeting, the audit system is tooth[less], is nothing, it’s just, being controlled”* County Manager 7, County B

Interview respondents reported that inflation of commodity prices caused inefficiency through stockout of commodities as elaborated below:

> *“… (inflated commodity prices) affect efficiency of services, because if you, if you, if you bring two items instead of four and inflate the price, because that those two have occupied, where you wanted to bring 100 gloves you bring 10, then the person will not be served promptly, and you have delayed service delivery.”* County Manager 4, County B

> *“…if you have diverted money from where it’s supposed to be and either a commodity or a service that was supposed to be delivered was not delivered, then you have interfered with efficiency.”* County Manager 4, County B

### Theft, Embezzlement and Misuse of Public Funds and Property

Theft of public property was reported. Examples included the theft of commodities such as medicines, fuel, and medical equipment by staff who would then sell these to the public.

> *“There are errand officers who are out also to pick items, machines; you know some machines are imported? Yeah and currently there is a machine that got lost in our ICU. And it is thirty million, we are still following up. The issue is at the director of criminal investigation’s office…if a machine for instance is stolen, a vital equipment is stolen, you see clients who are supposed to be served by that machine maybe take time before the hospital procures another one.”* County Manager 3, County A

> *“So, in some of our facilities, we have seen loss of items like nutrition commodities. In some cases, you find the watchmen were involved, that they had a key which nobody knew they had. Sometimes you look at those bill cards you find people have not balanced their bill cards. When you ask why, they say, they have too much workload, so not able to balance. But sometimes you get worried, because it could become a loophole for theft of commodities.”* County Manager 1, County B
>
> *“Sometimes our drivers collude with the pump attendants and not fuel the required amount, or even not fueling totally. For instance, yesterday, they were going to Eldoret, which is like 200 kilometres away. They are normally given 40 litres. For yesterday’s case, the pump manager called to ask me if we are fueling again yet we had fueled towards midnight[recently]…and from midnight the vehicle had not gone anywhere.”* County Manager 2, County A

It was also reported that there were cases of embezzlement of funds. This entailed the diversion of public funds meant for specific activities to individuals.

> *“…once in a while you will hear that facility funds have been embezzled, we take action, just like you see in Kenya everywhere, there are those individuals at all times, they are hell bent to do something, they will lay their hands on whatever is not meant for them.”* County Manager 2, County A
>
> *“I have a case where I have had to ask some staff to pay back (from their salaries) two hundred and eighty something thousand Kenya shillings to recover funds that they had embezzled in collusion the facility in-charge and the chairman of the health facility committee.”* County Manager 5, County B

Respondents reported that these forms of corruption were enabled by the collusion with and involvement of local political leaders, which weakened accountability.

> *“And we know there is this trend in Kenya that the big fishes, if you have money and you have a godfather, nobody can do anything to you…the much they can do is just transferring you to another pool, to another forest. So, it is not that pertinent or lenient punishment for these people.”* County Manager 1, County B

Interview respondents reported that corruption through embezzlement of funds affected both human resources for health and health service delivery. Service delivery was affected when funds were redirected from initially intended functions.

> *“But if you create bureaucracy, I think you get an opportunity because uh, depending on eh, who you are, if you have no patience and you are asked to pay 200,000 for you to get 500,000, then the best thing for you is to pay 200,000 and get 500,000 and do your work. But you see, you have already lost. At the end of the day, eh, eh, the results you are going to achieve is not as part of the budget allocated, because, uh, if it is a construction, the quality of that construction is going to be undermined.”* County Manager 7, County B

The compromise of service delivery, and inability to act to stop these forms of corruption led to demotivation of health workers.

> *“If you have not paid your casual let’s say your cleaners, your, your subordinate staff for X number of months, because you have, you have diverted the money or you have not, you are not willing to pay then they will be, you know, be a strike or maybe a go slow or something and then you have affected you know, affected how services are being done promptly…”* County Manager 4, County B

Interview respondents also reported cases of misuse of public property and equipment by staff. For instance, an interviewee in County B mentioned that public health officers used motorcycles allocated to them for their private use. It was reported that the failure by the county to maintain equipment, resulting in staff using their personal resources to maintain them was partly used as justification for the private use of public property.

> *“…we are having motorbikes which we’re using. And we have never come across that (as) misuse of the motorbikes. The maintenance sometimes relies on an individual because we may inquire for the maintenance, and we may not get the services. So, you end up using your own money to maintain the motorbike.”* County Manager 1, County B

### Informal Payments

Some interview respondents reported that some health workers would solicit for bribes from a patient with a huge bill to help them evade their responsibility of paying user fees. The quote below elaborates this further:

> *“For example, a patient is admitted in a ward, she stays for three/four days, is discharged but hasn’t paid. A rogue staff comes and says, “Umekaa, kwa nini shida ni nini?” (You’ve overstayed, what’s the issue?). “Bill nilipata ni 15,000 na sina.” (My bill is 15,000 but I can’t afford) So the rogue officer then says, “Kama ni 15, ambia watu wako watafute hata kama ni sita nione vile nitakusaidia (If your bill is 15 thousand shillings, ask your relatives to pay me at least 6 thousand shillings, then I will see what I can do to facilitate your discharge).”* County Manager 7, County B

The collusion between health workers and patients to avoid paying user fees led to loss of revenues by health facilities. A reduction in the amount of revenue collected by a facility in each quarter also reduced the budget that a facility could plan for in the following quarter.

> *“If we are to buy less drugs for patients, at the end of the day, in that quarter you may discover at one point you don’t have drugs. So, you’re prescribing for patients to go and buy (outside the hospital). So, the money you’d have received for the drugs will be paid in the chemist outside the hospital. So, the hospital will actually lose.”* County Manager 7, County A

Informal payments also presented a financial barrier to patients, reducing their access to healthcare services.

## DISCUSSION

We set out to explore the relationship between corruption and county health systems efficiency in one efficient and one inefficient county. We found that corruption practices were generally cross-cutting, with no clear distinction between the efficient and inefficient county. This finding could be because the nature of corruption practices documented are pervasive in Kenyan counties, with differences in degrees across countries that are difficult to tease out using a qualitative approach. It could also be because the counties that were ranked as efficient by the quantitative analysis by being on the efficiency frontier are inefficient in absolute terms, even though they are relatively more efficient than the counties that are at a distance from the efficiency frontier. This notwithstanding, the study found that corruption practices could potentially influence the efficiency of county health system in several ways.

First, corruption practices lead to inefficiency by causing a direct loss of healthcare resources because of theft, embezzlement and misuse of public resource and property. These forms of corruption have been reported as common in other settings. For instance, a review of corruption practices in the Nigerian health sector reported the diversion and private selling of health commodities procured by the public sector, and embezzlement of healthcare funds (19). Second, corruption practices led to an increase in the operational cost of health systems. This was because of inflated procurement costs perpetrated by collusion between county staff and private suppliers. Similar findings have been reported in South Africa where the overpricing of sub-standard products during procurement was reported (30). Third, corruption practices led to a reduction in the productivity of human resources for health. The reduced productivity of human resource for health was because of the politically influenced recruitment process that yielded less-than-competent staff. The influence of staff recruitment by political patronage has been reported in other settings (31). Reduced health worker productivity also arose because of reduced motivation resulting from the favouritism associated with recruitment and the allocation of training opportunities. Fourth, corruption practices led to the reduction in the quality of care delivered by health systems through the supply of sub-standard healthcare commodities which came about from collusion between suppliers and county staff. The supply of sub-standard health commodities due to corruption has been reported elsewhere. For instance, a study of corruption in the South African health system reported the supply of substandard surgical beds that broke down during surgery leading to the fracturing of a patient’s skull (30). Lastly, access barriers such as informal payments may compromise the delivery and coverage of healthcare services.

This study has several limitations. First, the study was conducted in 2 out of the 47 counties in Kenya, and hence not generalizable to all counties. As a qualitative study, it did not aim for statistical generalizability, but rather for analytical generalizability. Second, given that corruption is a sensitive and difficult topic to discuss, it is likely that social desirability bias affected the responses of study participants. We found that interview respondents were cautious when sharing their perceptions about corruption in the health system. They avoided giving specific examples or examples closer to their work settings, preferring instead to share information in general terms, and providing examples outside their specific work settings (e.g., other departments or health facilities, rather than the ones they were stationed). Similar challenges with willingness of study respondents to provide information on corruption have been reported by a study that examined health sector corruption in Nigeria (32). However, we found that it was easier to obtain information from respondents who had either been in the health system for longer, or influential persons in senior positions. A possible explanation for the easier cooperation of more senior and/or experienced respondents is that they felt less vulnerable to victimization from sharing information on corruption. Lastly, the findings of the study would have been strengthened if the study included an examination of the existence, strengths, and weaknesses of anti-corruption strategies in place in the county health system. This would have informed the strengthening and/or introduction of specific anticorruption strategies to tackle the existing corrupt practices at the county level of the health system. There is however relatively little evidence about strategies to address corruption in general (33).

## CONCLUSION

This study reports the existence and nature of corruption practices in the Kenyan health system at the county level. It further highlights the link between these corruption practices and the efficiency of county health systems. Addressing these corruption practices is a likely pathway to enhancing the efficiency of county health systems in Kenya. Further studies are required to explore the feasibility and effectiveness of anti-corruption practices to address the observed corruption practices in the Kenyan health sector.

## LIST OF ABBREVIATIONS

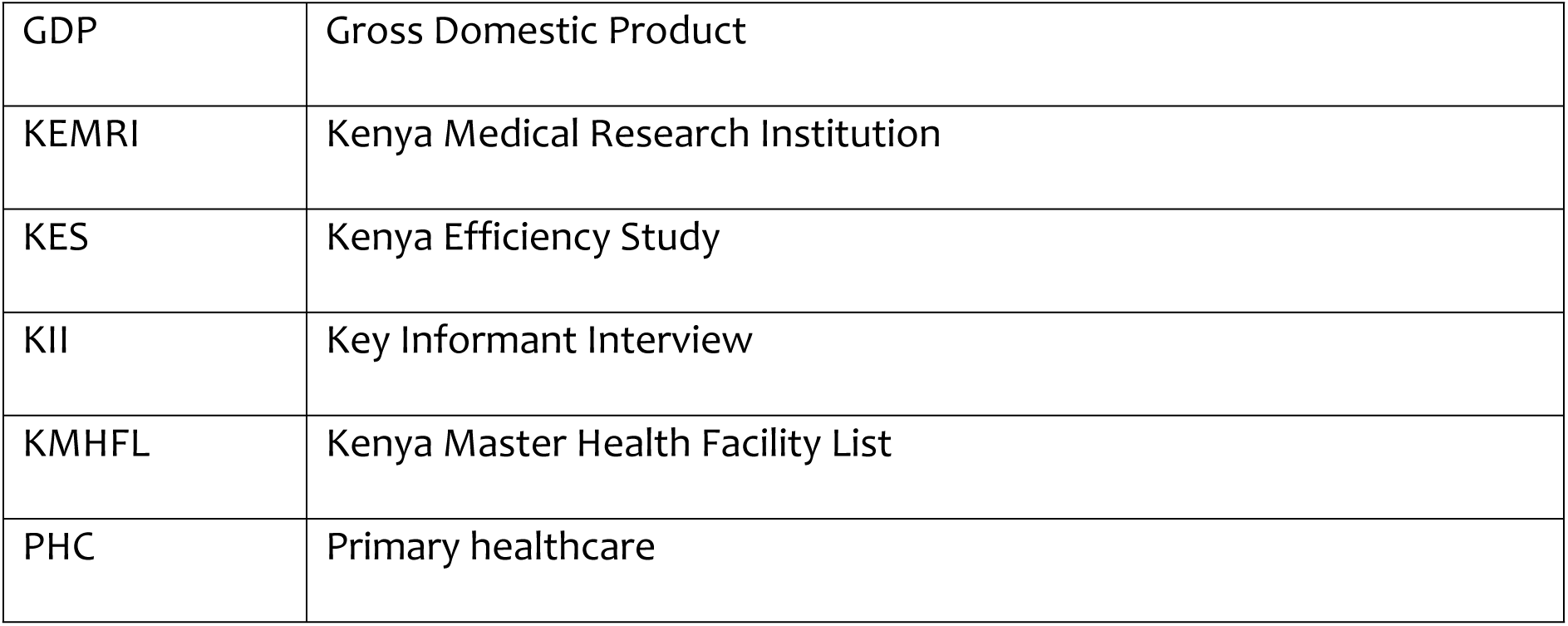
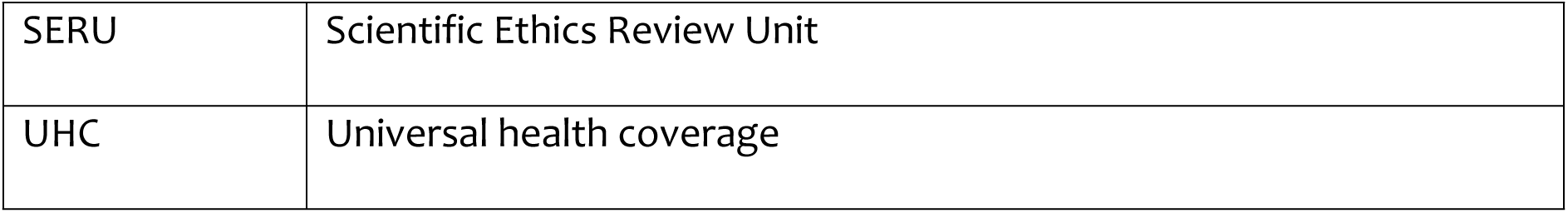

|  |  |
| --- | --- |
| GDP | Gross Domestic Product |
| KEMRI | Kenya Medical Research Institution |
| KES | Kenya Efficiency Study |
| KII | Key Informant Interview |
| KMHFL | Kenya Master Health Facility List |
| PHC | Primary healthcare |
| SERU | Scientific Ethics Review Unit |
| UHC | Universal health coverage |

## DECLARATIONS

### Ethics approval and consent to participate

Ethics review and approval (KEMRI/SERU/CGMR-C/154/3814) for this study was obtained from the KEMRI Scientific and Ethics Review Unit (SERU). Approvals from other relevant authorities were also sought prior to commencement of the study. All methods were performed in accordance with the relevant guidelines and regulations. All study participants provided written informed consent to participating in the study and being audio recorded. Confidentiality was assured by anonymizing the study counties, de— identifying respondent data, securing the collected data in password protected computers, and restricting access to the data to researchers only.

### Consent for publication

Not applicable.

### Availability of data and materials

The datasets generated and/or analysed during the current study are not publicly available due to participant confidentiality but are available from the corresponding author [EB] on reasonable request.

### Authors’ contributions

Conceptualization of the study was done by Edwine Barasa, Julie Jemutai, Kara Hanson, Sassy Molyneux, Charles Normand, and Benjamin Tsofa. Data collection was done by Joshua Munywoki, Lizah Nyawira, Anita Musiega, and Rebecca Gathoni. Formal analysis was carried out by Joshua Munywoki. Joshua Munywoki drafted the initial manuscript which was subsequently revised for important intellectual content by all authors. All authors read and approved the final manuscript.

## Acknowledgements

We acknowledge all the health system stakeholders from the national and county level, as well as non-state actors who participated in this study.

